# Modelling the risk factors of malnutrition among the children below five years of age in Uganda: A GSEM-based analysis

**DOI:** 10.1101/2022.04.18.22273977

**Authors:** Vallence Ngabo M, Leonard Atuhaire, Peter Clever Rutayisire

## Abstract

This study aimed at establishing the causal mechanism through which the risk factors of malnutrition act to influence the undernutrition of under-fives in Uganda. A generalized structural equation model (GSEM) was estimated based on the multifaceted nature of the risk factors according to the UNICEF 1991 conceptual framework for child malnutrition. Uganda Demographic and Health Survey (UDHS-KR) dataset was used for the analysis. An index of malnutrition which was the overall response variable in the GSEM model was derived from the three anthropometric variables of stunting, underweight, and body wasting.

The findings indicate that sex of the child, age of the child, the perceived size of the child at birth, the weight of the child at birth, duration of breastfeeding, mother’s education level, working status of the mother, wealth quintile of the household and mothers’ weight (BMI) were the factors that had a direct and significant influence on undernutrition. Further analysis of the causal mechanism reveals about five main paths of causal mechanism and these were; (i) the mother’s level of education influences the mother’s weight which influences the birth weight of the child and then this combination influences under-nutrition, (ii) mother’s education levels also influence the household wealth status and as observed above, household wealth significantly influences under-nutrition, (iii) residence whether rural or urban setting influences under-nutrition through its influence on the household wealth, (iv) being in urban areas is also associated with employment opportunities which in turn influences the duration of breast feeding and then influences under-nutrition, (v) size at birth which is key in influencing under-nutrition is influenced by the working status of the mother which is dependent on the place of residence.

Based on the findings, the recommendation is that a multi-faceted approach that addresses all the factors at different levels be implemented in order to get a lasting solution to the silent killer of under-fives.

## Introduction

Malnutrition, in all its forms, includes undernutrition (wasting, stunting, and underweight), inadequate vitamins or minerals and over nutrition which includes overweight, obesity, and resulting diet-related non communicable diseases. World over about 45.4 million children under 5 years of age (6.7%) are wasted, 14.3 million are underweight while about 149.2 million (22%) are stunted (1). Undernutrition is the most worrying nature of malnutrition and is linked to about 45% of deaths among children under 5 years of age worldwide (2). The developmental, economic, social, and medical impacts of the global burden of this health condition are serious and lasting, for individuals and their families, for communities and for countries while the victims of undernutrition have higher chances of mortality.

The dangers of malnutrition are long lasting and they may go beyond the physical impairments of the malnourished children to retarding their cognitive and educational achievements (3,4) correlates anthropometric measures and undernourishment and established that undernourishment of children in their early years was responsible for lowering children’s non-verbal IQ significantly. Early childhood illness like diarrhoea coupled with malnutrition influences cognitive impairment in later childhood(5–7). Many other studies (8–11) all appreciate the fact that malnutrition is an important influencer of cognitive and education performance of children and of course later productivity of individuals.

Malnutrition is influenced by different factors differently in different regions, economies or societies. Major crime rate which was an indicative of reduced income in the society was found to be more significant in influencing malnutrition than poverty index or low educational attainment in Jamaica (12). In Guatemala, the significant predictors of child malnutrition were related to the health status of the child, number of children <5yrs old and the household size (13). Maternal malnutrition and urban concentration of households were found as the most significant risk factors of child malnutrition among the tribal children in India (14). This further confirms the fact that maternal malnutrition influences the health status of the mother in terms of size and weight (body mass index) which later feeds into child health status.

Uganda, just like many other least developed countries (LDCs) especially in Africa, still experiences an unacceptably high rate of malnutrition. With a national level prevalence of about 3 in every 10 children (2.4 million) of all children below 5years of age stunted, more than 10% and more than 4% of the children below five years of age having defects of underweight and body-wasting Ugandan children are among the most victimized children in African and the world at large (15). Bearing in mind the consequences of malnutrition, multi-sectoral interventions with a goal to improve nutrition of mothers and children (16) were designed and implemented but the fact is that after 5 years and by the end of 2016, Uganda still had high and unacceptable prevalence rates.

Many studies on malnutrition in Uganda as cited in the literature, considered the problem with little attention to the mechanism, pathways and structural system of causation (17– 22). The structural mechanism of operation needs to be given much attention because the malnutrition problem is multifaceted and complex in nature. Paying attention to the general nature, confounding, mediating and multilevel nature of factors where some factors are simultaneously endogenous and exogenous guarantees a deeper understanding of the core causes and structural mechanism of malnutrition. Furthermore, the dynamics of the economy implore a continued examination of the malnutrition prevalence and its associated risk factors using appropriate and advanced statistical methods. The mix of variables that influence malnutrition encompasses all nature of variable such as; count, continuous, discrete and categorical. The way in which these factors operate to influence malnutrition is complex and multifaceted whereby some variables feed into each other in a structural way which making structural equation modeling an appropriate approach. Indeed, since the mix of variables involves count, discrete, continuous and categorical variables and since the system of equations involves more than one response variables that must be estimated simultaneously, structural equation modelling process in its generalized form (GSEM) appears to be the most appropriate model. GSEM has the power to analyze a mix of count, continuous and categorical data while accommodating their hierarchical nature as well (23). As such, the current study exploited structural equation models (SEM) in its generalized form (GSEM) (24,25). This helped in gaining a deeper understanding of the causes of the persistent malnutrition problem as we search for a long-lasting solution to the leading cause of child morbidity and mortality in Uganda and world at large.

## Methods and materials

The current investigation was based on secondary data that was extracted from the Uganda Demographic and Health Survey (UDHS-2016). For the purpose of this investigation which considers the children below five years of age, the extraction considered the data from kids record (KR) files which contains the data for children age 0-59 months alongside their respective attributes such as the socioeconomic, demographic, and child level factors. The survey was designed to provide estimates of the population and health indicators including fertility and child health for the country as a whole for rural and urban separately and for each of the 15 regions in Uganda (South Central, North Central, Busoga, Kampala, Lango, Acholi, Tooro, Bunyoro, Bukedi, Bugisu, Karamoja, Teso, Kigezi, Ankole, and West Nile). Different units of analysis were taken into account including; household, individual, children age 0-5 years, women aged 15-49 years and men aged 15-54 years.

The 2016 UDHS survey was keenly carried out by an experienced team that managed to collect and process data for 29,226 children aged between 0 to 59 months and successfully recorded anthropometric measurements for about 4,530 children. More to the anthropometric variables, data was collected on several other variables such as household related characteristics and socioeconomic variables. The over-all dependent variable was malnutrition-status of the child which was binary in nature. Using principal component analysis, the status of child malnutrition was derived from the three anthropometric indices of stunting, under-weight and wasting. Having found a high correlation coefficient between the three anthropometric indices, it necessitated the creation of a single composite index from the respective indices of stunting, underweight and body wasting (26). The first principal component was used to derive a new index of malnutrition which was later used to classify the nutrition status of children using *zscores as* per World Health Organization child growth standards of 2006. The z-scores (*Zi*) are calculated as 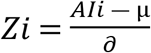 where Ali refer to the anthropometric indicator of i^th^ child, µ *and ∂* respectively refer to the median and the standard deviation of the reference population. All children whose z-scores were below -2SD from the median or reference population were categorised as malnourished while all children whose z-scores were equal or above -2SD from the reference median were categorized as normal. This led to generation of a derived proxy variable (malnutrition status) that was categorical and binary (equation 1).

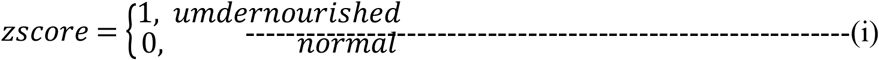

More other variables were derived form the dataset and used in the modeling process. Variables such as mother’s body mass index were derived from mother’s weight using the z-score for the mother’s weight and the resultant variable (MBMI) was categorized as thin (MBMI ≤-2SD), normal (−2≤MBMI≤2SD) and obese (MBMI≥2SD). Other variables as well as their hypothesized mode of interaction are represented in the conceptual framework. The risk factors were categorized into four; the distal factors, the intermediate factors, the immediate factors and the child inherent factors. With GSEM modeling approach, variables at some levels feed into each other’s to bring out the overall effect as seen in Fig 1

**Figure 1.**
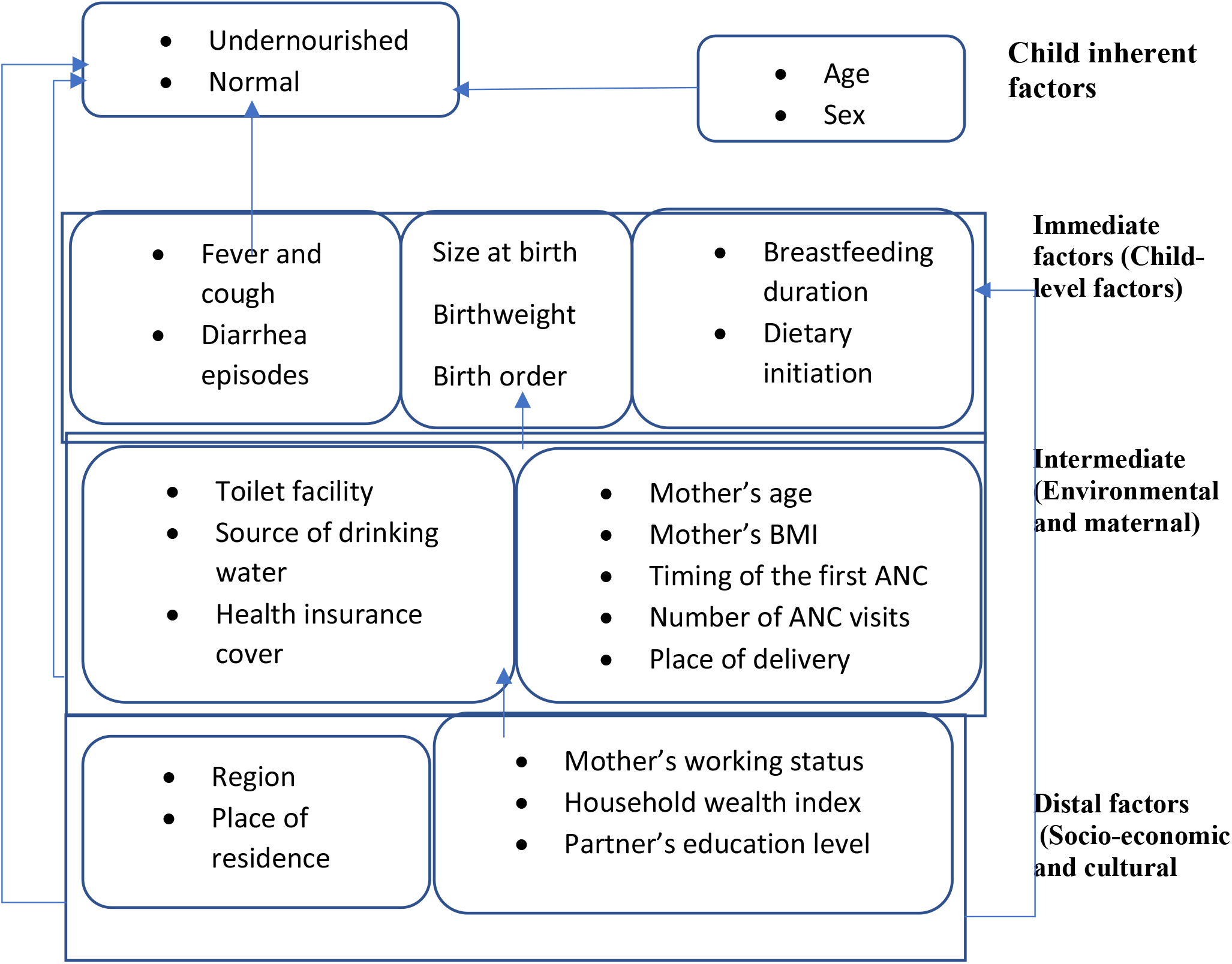
A conceptual framework for analyzing the risk factors of under-five Undernutrition.

The hierarchical and interrelated nature of risk factors of under-five malnutrition calls for a model that incorporates and can estimate a system of equations simultaneously. A system that considers several nodes and as such each end node where variables feed into each other be treated as a dependent or response variable as represented in the conceptual framework (Fig 1). In such a system, several response variables do exist. Consideration was also given to the statistical nature of variables, that is whether the variables were quantitative or qualitative, their scale of measurement and their statistical distribution (link and family). As such, a GSEM was considered for this study since it meets these criteria.

GSEM is a family of statistical techniques that is suitable for the analysis of multivariate, multi-level, categorical, and ordinal data. It allows in the same model, the analysis of a wide range of variable responses. It’s very vital in the analysis of complex linkages of variables in models where variables are either exogenous, endogenous or both. The effects of variables in the complex structure of the model are measured or determined simultaneously. Generalizations is important in understanding the structural and simultaneous nature of the interaction of variables in malnutrition problem. GSEMs represents a generalization of SEMs and has an advantage of accommodating all types of variables by considering their link and family distributions (27). GSEM also has other advantages over other modeling approaches such as; it relaxes the assumption of normality of variables, it allows for multi-level or hierarchical modeling and it allows for group analysis.

More so, GSEM combine the power and flexibility of both structural equation models (SEM) and generalized linear model (GLM) in a unified modeling framework (28). A combination of the statistical distribution of the family (*F*)and the link function g (·) for the response variables was clearly specified within the structural framework. Since undernutrition was a binary variable coded as 0 for a normal child and 1 for a malnourished child. A GSEM of Bernoulli family with the link function (Probit) was applied to analyze the risk factors to the malnutrition status of under-fives which is undernutrition in this case.

Therefore, in the current study the linear model of the format *yi* = *β′xi*+ *ui*, where *x* is a vector of exogenous variables was generalized to the linear model of the form *g*{*E*(*yi*)} = *β′xi* with *yi* ∼ *F*. The idea of family and link function was specified for endogenous variables of every equation in the system of model (Fig 2). Sampling weight was used to account for disproportionality in the samples caused by survey design. STATA version 15 was used for the analysis and results were judged at 95% confidence level.

**Figure 2:**
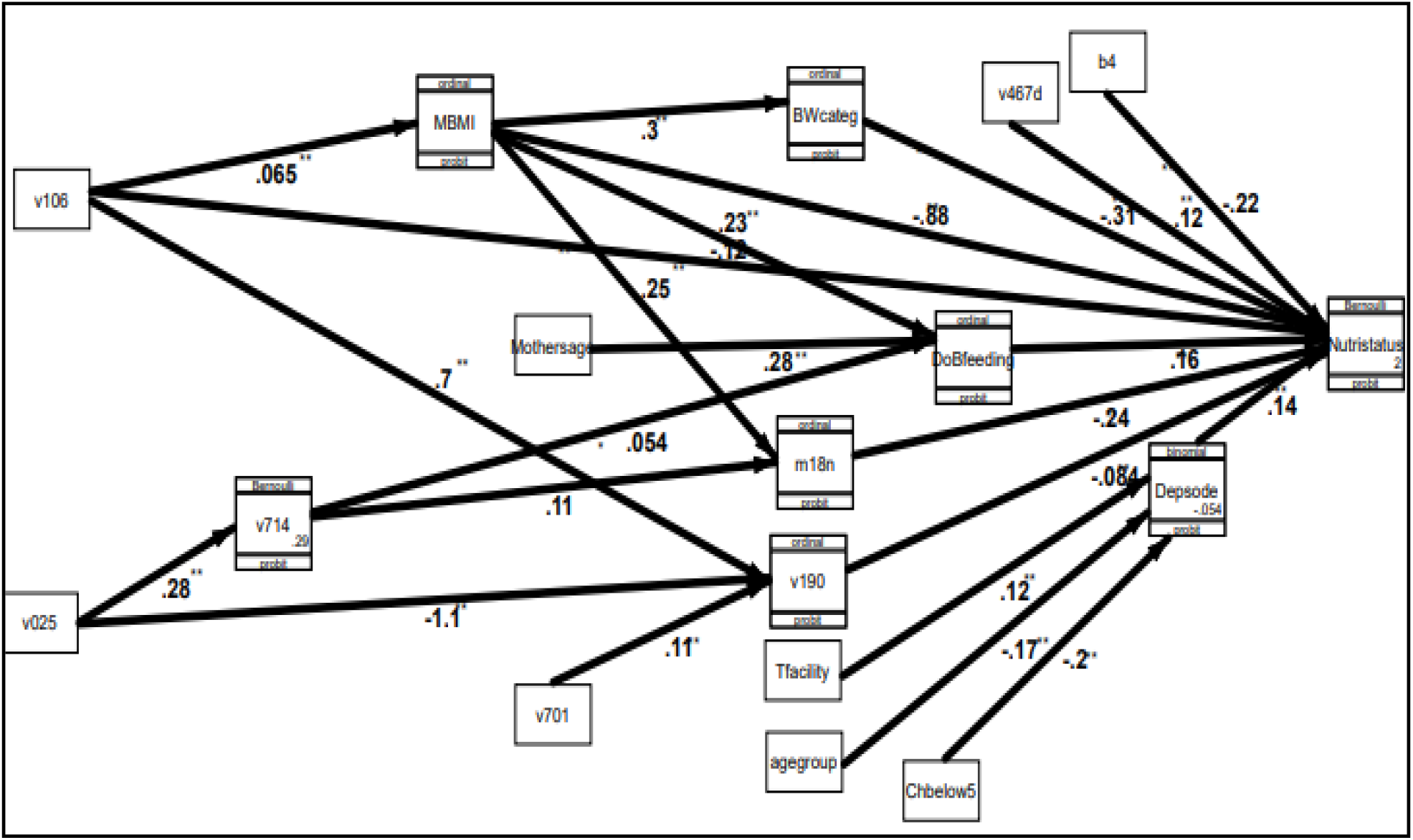
The generalized structural equation model showing the causal mechanism. Observations made from Fig 2 indicate that, among other factors that play a role in the causal mechanism are; father’s education level which has a big influence on the household wealth, a combination of toilet facility, age group of the child and the number of children below 5 years of age in the same household influences under-nutrition through their effect on diarrhoea episodes. duration of breastfeeding is influenced by the mother’s age, mother’s working status and mother’s weight which are in turn dependent on residence (v025) and mother’s education level (v106). The household wealth index influences undernutrition and is itself dependent on where the household resides (rural/urban), the education levels of both the mother of the child and her partner. The weight of the child at birth which was key in influencing under-nutrition negatively is itself dependent on mother’s weight which also depends on the mother’s education level.

### Ethics statement

This paper is based on secondary data that exists and is publicly accessible upon request. Accordingly, the permission to use the UDHS 2016 dataset was requested via the DHS website (https://dhsprogram.com/data/dataset/Uganda_Standard-DHS_2016.cfm?flag=1). The DHS programme is a worldwide reputable organization for collecting and disseminating credible national representative data on health indicators especially data on fertility, family planning, maternal, child health, HIV, malaria and nutrition. On behalf of the country team, the ICF institutional review board (IRB) reviewed and approved the UDHS 2016 tools for the survey as well as the procedure of obtaining written informed concent. The ICF IRB complied with the United States Department of Health and Human Services regulations for human subjects. Informed consent was obtained from participants in writing whereby a form was signed by the respondent as well, data on minors were collected on children based on mothers’ informed consent obtained in writing. Participants were given full information about the survey and their participation was purely voluntary. Confidentiality and keeping participants anonymous, elements were observed by exclusion of identifiers from the final dataset obtained. Further information regarding the conduct of the study that generated the data used can be found in the report of UDHS 2016 (29).

## Results

The interrelationship that is depicted by the conceptual framework (Fig 2) dictates estimation of both the direct and indirect effects of the risk factors on undernutrition in order to establish the path and causal mechanism of the risk factors (Fig 2). Most variables were used as they were named in UDHS database while other variables such as the z-scores and malnutrition index were derived from the ones in the database. As such, there was a need to harmonise the dataset variables and the narrative whereby; b4 (sex), bord (birth order), m18 (size at birth), h11 (had fever episodes), h22 (had malaria episodes) v012 (mothers current age), v106 (mothers education level), v701 (partners education level) v481 (insurance cover), bwgt (birth weight), v190 (wealth quintile) and v025 (residence). The results were generated and discussed at three level, the direct effect of combined factors, the effect at factors levels and then the paths analysis which demonstrated the mechanisms for the significant factors at level one and two. With guidance from the conceptual framework, variables included at any level had a possibility of influencing undernutrition either directly or indirectly through mediating variables. Judgement was made at 10% level of significance (90% confidence interval) and a variable was thus considered a statistically significant influencer of undernutrition whenever its p-value (p(z)) was less than or equal to 0.1. It is important to note that, analysis was modified with sampling weight to correct for the disproportionality of the sample during the survey and accordingly, the women’s individual sampling weight (v005) was used. Table 1 presents the results of analysis of direct effects.

**Table 1:**
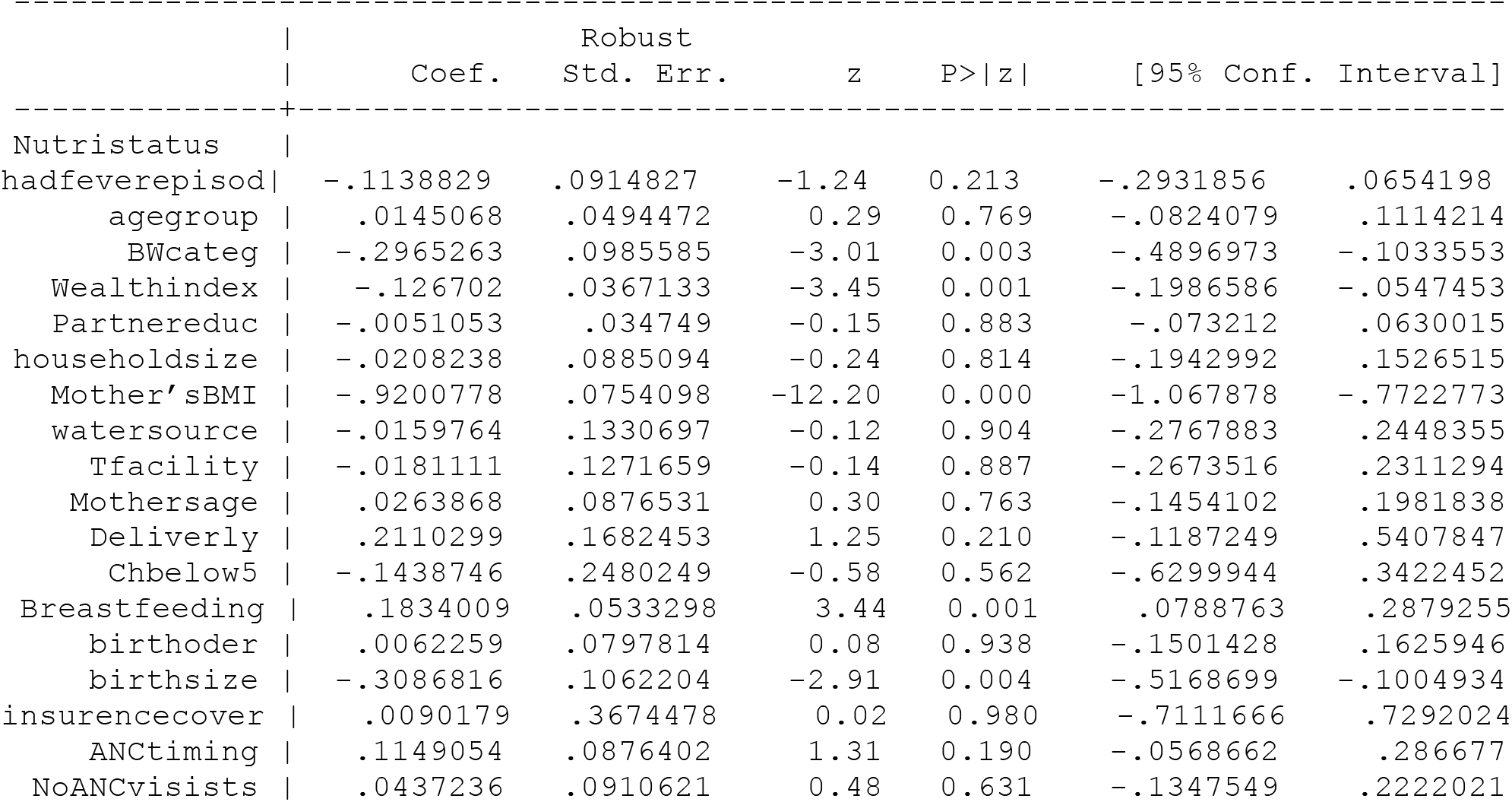
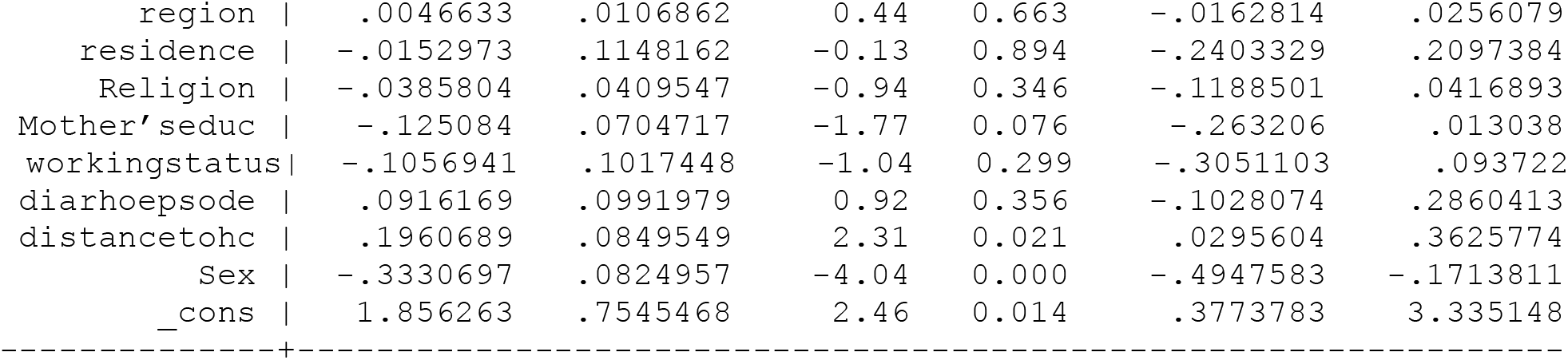
Direct determinants of undernutrition for children 0-59 months (n=1,731)

A total of 4530 observations were entered into the GSEM model but due to missing values, only 1,731 observations were successfully considered for the analysis. From table 1, it can be observed that the distal or socioeconomic factors that significantly influences undernutrition were; mothers’ level of education combined and the combined household wealth index. The most significant influencers at intermediate level (environmental and maternal factors) were distance to the health center and mothers’ weight which was proxied by mother’s body mass index. Child level factors also which are well known as child level factors; duration of breast feeding (DoBfeeding), perceived size at birth (m18n) and weight at birth (BWcateg) were the most statistically significant influencers of undernutrition at child level. Among the child inherent factors, only sex of the child turned out to be statistically significant in influencing undernutrition of under-fives in Uganda.

The study also considered analysis at factors level with an aim of identifying which factor levels among the categorical variable were responsible for the effect. It also observed that, there was a possibility for variables which were insignificant in combinations to have some factor levels within themselves that turned out statistically significant. Table 2 shows the results of how risk factors directly influenced under-nutrition at factor levels.

**Table 2:**
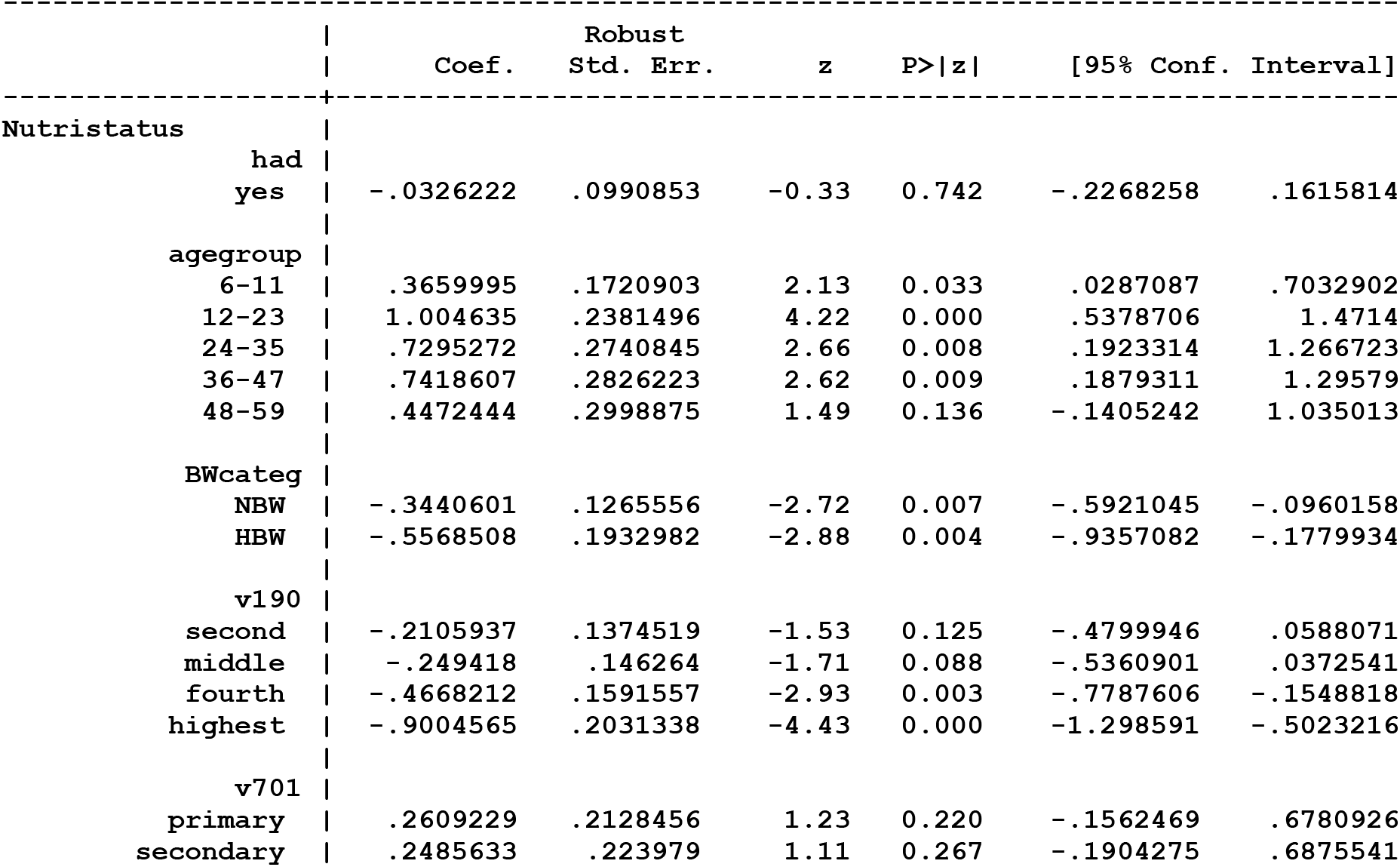
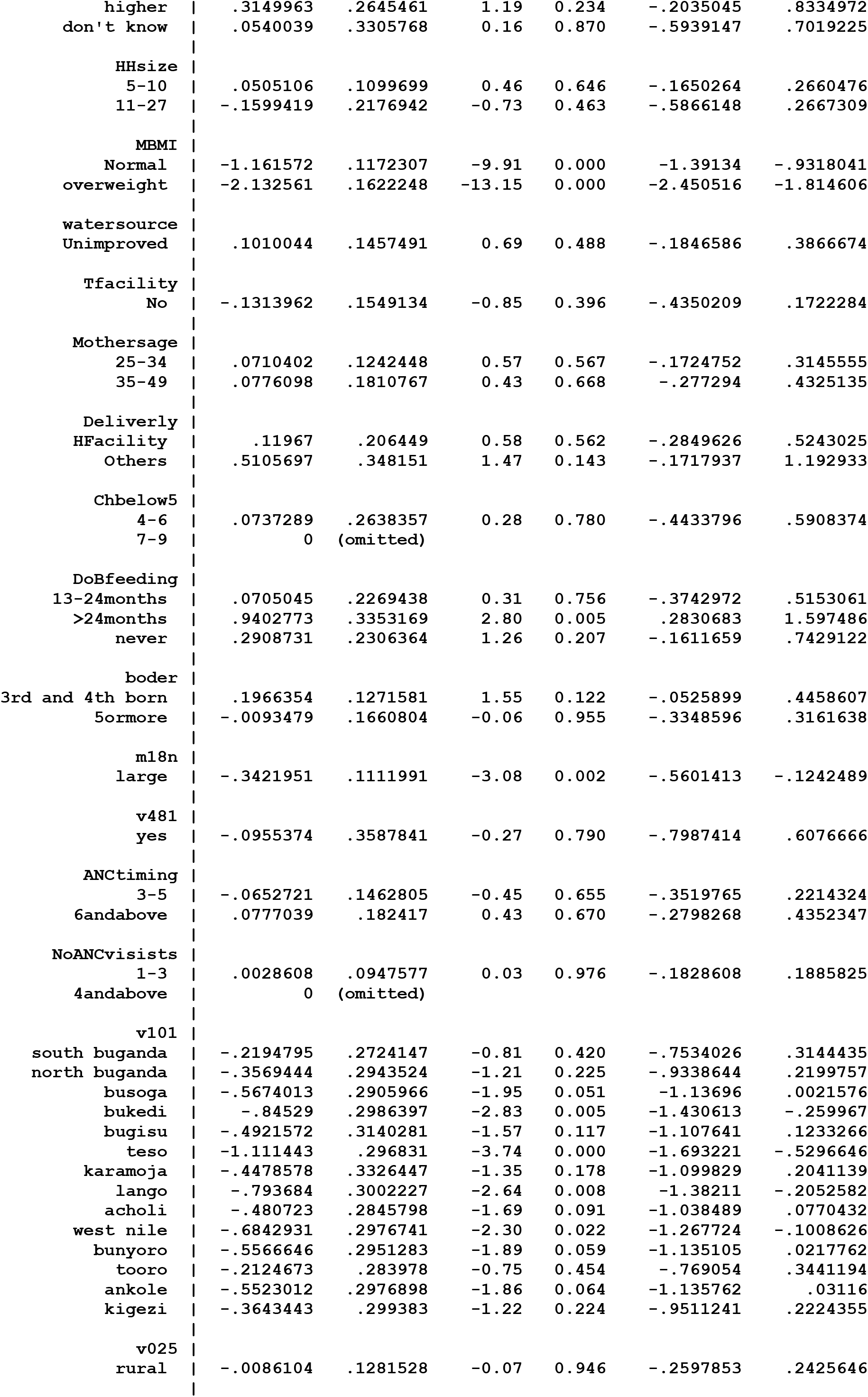
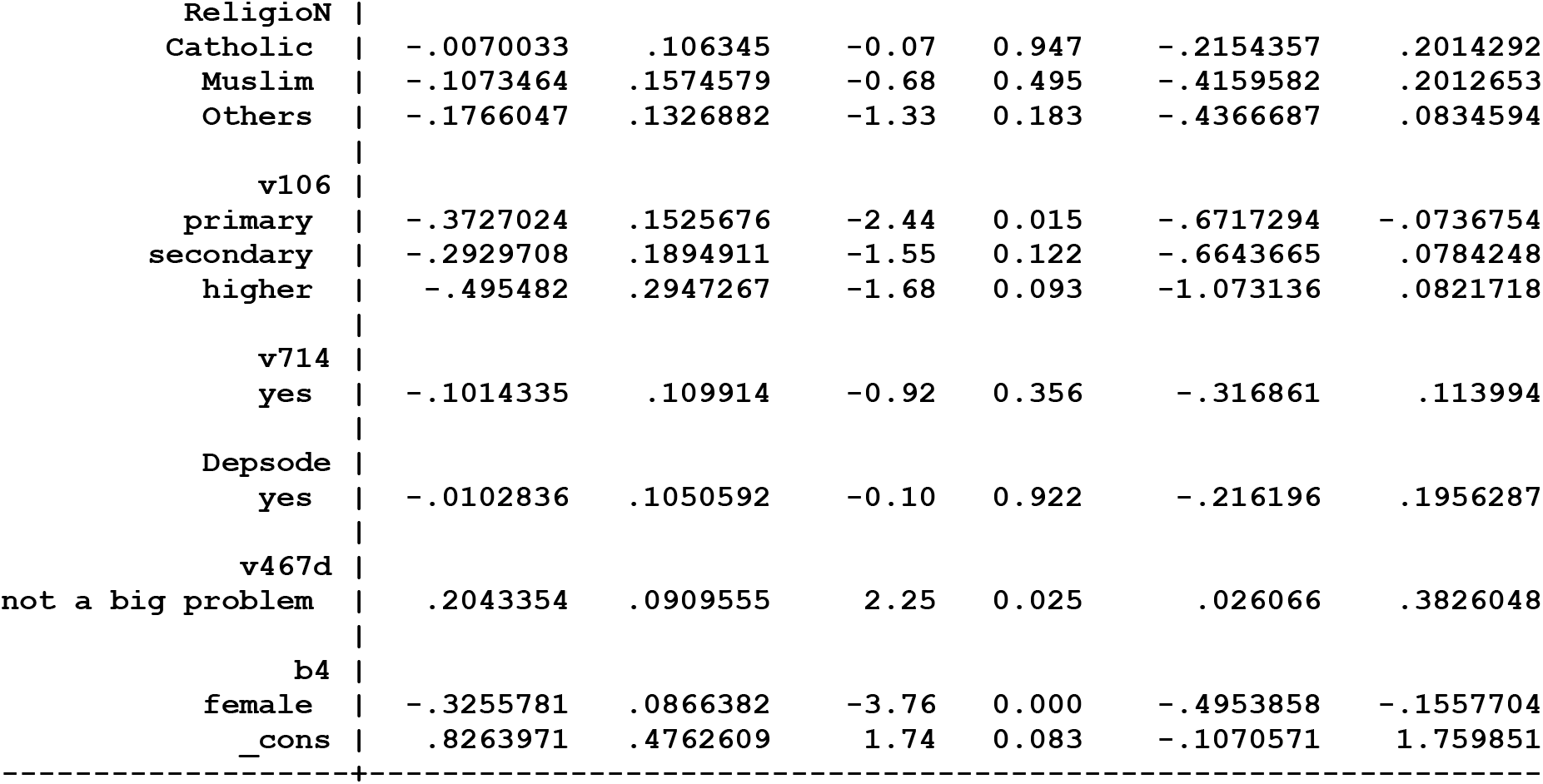
Direct determinants of undernutrition for children 0-59 months at factor level (n=1,731)

Reference made to Table 2 indicates that being a female child is significantly different from being a male child as it can clearly be seen that the female sex is negatively associated with under-nutrition of children below five years of age in Uganda as opposed to their male counterparts. Being nearer to the health center (v467d) has a positive effect on under-nutrition and the effect is significantly different from the effect of short distances to health centers. Mother’s level of education beyond primary is negatively associated with under-nutrition and the difference is significant from mother’s with less than primary level of education though the difference between mothers with secondary level of education and mothers with no formal education isn’t statistically significant. Region wise, Ankole, Busoga, Lango, Westnile, Acholi, Bukedi and Bunyoro were statistically different from Kampala while, Kigezi, Tooro, Karamoja, South and North Buganda were not statistically different from Kampala region as far as being associated with under-nutrition is concerned. The perceived size of a child at the time of birth negatively and significantly influences under-nutrition for children whose size was perceived as large and as opposed to those children whose size at birth was perceived as small. Mother’s perception of the size of the child at birth is subjective and somehow ambiguous since it has no standard scale and as such this outcome may be cautiously consumed. It can be observed that breastfeeding of children beyond two years influences under-nutrition positively and this outcome is statistically different from the base category of breastfeeding between 0-12 months. The category of breastfeeding between 13-24 months just as those who had never breastfed was not significantly form the base category. Body weight as proxied by mother’s body mass index (MBMI) significantly influences under-nutrition positively for thin mothers as opposed to the normal and overweight mothers. The fourth and fifth wealth quintiles negatively influences under-nutrition and the influence was significantly different from wealth quintile one. The second wealth quintiles were negatively associated with under-nutrition though it was not significantly different from wealth first wealth quintile. Birth weight category was significant and negatively associated with undernutrition. Being born with normal birth weight and higher birth weight influences under-nutrition negatively and this is statistically different from being born underweight.

Deeper analysis commenced by looking at the causal mechanism of the risk factors of undernutrition. To avoid overcrowding of the path diagram, only factors that were directly significant in influencing under-nutrition were considered for this analysis. Accordingly, weight of the child at birth, duration of breast feeding, perceived size of the child at birth, sex of the child, mothers’ weight, diarrhoea episode, household wealth index and distance to the health center were analysis for influencing variables (Fig 2).

Briefly some of the possible identified causal mechanism of under-nutrition may operate through;

i. Mother’s level of education influences the mother’s weight which influences the birth weight of the child and then this combination influences under-nutrition
ii. Mother’s education levels also influence the household wealth status and as observed above, household wealth significantly influences under-nutrition
iii. Residence whether rural or urban setting influences under-nutrition through its influence on the household wealth
iv. Being in urban areas is also associated with employment opportunities which in turn influences the duration of breast feeding and as seen in the previous section influences under-nutrition
v. Size at birth which is key in influencing under-nutrition is influenced by the working status of the mother which is dependent on the place of residence.
vi. The rest of the factors; sex of the child and distance to the health center are purely exogenous in this model.

It was observed that in this causal mechanism, the age of the child which is proxied as agegroup influenced under-nutrition through its effect on diarrhoea episodes alongside the nature of toilet facility (Tfacility) and the number of children below five years in the household.

## Discussion of the results

The main objective of the study was to identify the paths through which the major risk factors of under-fives malnutrition in Uganda. A GSEM (Fig 2) was proposed and consequently estimated. The risk factors were hierarchical in nature and thus fed into each other and finally influencing malnutrition outcome either directly or indirectly through mediator variables. From the estimated model, the most influential factors significantly and directly associated with under-five malnutrition of in Uganda as discussed in section 3 above were; sex of the child, child’s age group, birth order of the child, perceived size at birth, birthweight, breast feeding less than 12 months, wealth quintile, mother’s working status, mother’s body mass index (BMI), mother’s education level, partner’s education level, residence and region. Further exploration of the mechanism of how the risk factors influence malnutrition status involved establishing the factors behind them apart at each level. Region, sex and age-of-child were the risk factors that theoretically don’t have background influencers.

Being born male is positively associated with the risk of being malnourished. Earlier studies as well indicate that the probability of being malnourished was found significantly lower for females as compared to male children (30–32). The child age or age group and the duration of breastfeeding are some of other factor of concern in the drive towards a better child nutritional status. The duration of breastfeeding is an aspect of getting the body ready to absorb food nutrients other than breastmilk. Exclusive breastfeeding is necessary in the first five months as it is associated with better growth and care must be observed when introducing complementary feeding (33). The age of weaning is key in reducing malnutrition (34) while malnutrition increases with an increase in age of the child (35). In line with the current findings, the size and weight of the child at birth are other key factors that are significant in influencing malnutrition status world over (36– 38). The association between the mother’s weight (BMI) and age at first birth and child’s malnutrition status is evident in the current study and this fact was evident in other countries (39– 42) though sometimes malnutrition was found within households with obese mothers(43–45). The Ugandan setup is such that, the rate of increase in dependency may not much the rate of facilitation especially care and food security(46) while the number of children below five years in a household is a significant influencer of malnutrition (47). The issue of food insecurity and number of children below five years in a household which normally implies increased dependency which coincides with the rank of child’s birth as the current findings affirm that the third born are the main victims of malnutrition in a household.

Parents education important in appreciating the importance of childcare and nutrition. The current study reveals the great association between mothers’ formal education level and healthy children. Mothers with less than formal primary education were associated malnourished children while the reverse was true for the partners formal level of education. This shows that malnutrition can best be fought by putting more emphasis on teaching mothers with no formal education and emphasizing girl child education(42). On the same notes education of the girl child and mothers improves their employability skills, increases the odds of getting employment, earning and self-reliance in upkeep and child care practices but in most countries, mothers employment tends to increase malnutrition of under-fives (48,49) which calls for policies to consider mothers especially of younger children as a special category of workers at workplaces. Increasing employment opportunities for mothers therefore is key in reducing malnutrition of under-fives as the association between working status of mothers and nourished children is evident (50) but attention should be taken to improve mothers working and employment conditions (51). Poverty which is reflected by the wealth index is found to be a significant influencer of malnutrition, not only in Uganda (48). Malnutrition interventions should take into account the spatial factor of regional disparities due to the fact that different regions are affected differently (52–55) and the difference is obvious due to the fact that different regions are endowed differently for example income inequality between rural based and urban based regions is evidence (54) and social development (56).

Emphasis should be given to the path through which the risk factors malnutrition act. Improving the socioeconomic status like the education, wealth and employment opportunities of households (57) influences the health seeking behavior of women (58) and utilization of health care services (59,60) which in turn influences the health living of expectant mothers that influences the optimal weight and size of the baby at birth (61) that is key in reducing malnutrition among the under-fives (62).

## Conclusions

Malnutrition especially the under-nutrition of children below five years of age in Uganda is still a major public health problem that needs to be handled with great attention. The study reveals that factors that influence malnutrition are multi-faceted and complex and they feed into each other ranging from distal, intermediate, immediate, child level and child inherent factors. Nutritional interventions should take into consideration the paths and the interaction mechanism of the risk factors. The strategies and policies against malnutrition should be multifaceted and considerate of the interconnected or interrelated nature of the risk factors and policies should simultaneously consider all the factors along path(s).

## Data Availability

Data was accessed with permission from DHS programme and is available at https://dhsprogram.com/data/dataset/Uganda_Standard-DHS_2016.cfm?flag=1

## Acknowledgement

The authors are so grateful to their respective institutions of higher learning for providing an enabling environment and time to have this work in place. We are grateful to African Center of Excellence in Data Science (ACE-DS), School of Economics, Gikondo campus at University of Rwanda for providing and enabling environment. We also extend our sincere thanks to the DHS programme for granting us permission and access to the UDHS data which has made this manuscript possible.

## Authors contribution

**Conceptualization:** Vallence Ngabo M, Atuhaire Leonard and Rutayisire P. Clever

**Methodology:** Vallence Ngabo, Atuhaire Leonard, Peter Clever Rutayisire.

**Formal analysis:** Vallence Ngabo M

**Writing the draft:** Vallence Ngabo M

**Review:** Atuhaire Leonard, Peter Clever Rutayisire

**Editing:** Vallence Ngabo M

